# Epidemiological and clinical insights from SARS-CoV-2 RT-PCR cycle amplification values

**DOI:** 10.1101/2021.03.15.21253653

**Authors:** Samuel Alizon, Christian Selinger, Mircea T. Sofonea, Stéphanie Haim-Boukobza, Jean-Marc Giannoli, Laetitia Ninove, Sylvie Pillet, Thibault Vincent, Alexis de Rougemont, Camille Tumiotto, Morgane Solis, Robin Stephan, Céline Bressollette-Bodin, Maud Salmona, Anne-Sophie L’Honneur, Sylvie Behillil, Caroline Lefeuvre, Julia Dina, Sébastien Hantz, Cédric Hartard, David Veyer, Héloïse M Delagrèverie, Slim Fourati, Benoît Visseaux, Cécile Henquell, Bruno Lina, Vincent Foulongne, Sonia Burrel, SFM COVID-19 study group

## Abstract

The SARS-CoV-2 pandemic has led to an unprecedented daily use of molecular RT-PCR tests. These tests are interpreted qualitatively for diagnosis, and the relevance of the test result intensity, i.e. the number of amplification cycles (*C*_*t*_), is debated because of strong potential biases. We analyze a national database of tests performed on more than 2 million individuals between January and November 2020. Although we find *C*_*t*_ values to vary depending on the testing laboratory or the assay used, we detect strong significant trends with patient age, number of days after symptoms onset, or the state of the epidemic (the temporal reproduction number) at the time of the test. These results suggest that *C*_*t*_ values can be used to improve short-term predictions for epidemic surveillance.

---

Molecular testing is a key component of screening policies to control the spread of infectious diseases and the SARS-CoV-2 pandemic has led to an unprecedented testing rate using reverse transcription polymerase chain reaction (RT-PCR) assays. (*1*). In clinical and public health practices, RT-PCR results are qualitative for viral respiratory disease diagnostics, with reports such as ‘positive’, ‘negative’, ‘uninterpretable’, and, sometimes, ‘weakly positive’. These are based on the cycles threshold, also referred to as crossing point or crossing threshold (here denoted *C*_*t*_), which corresponds to the number of PCR amplification cycles required for the fluorescent signal to rise above a positive threshold. In theory, the more abundant the genetic target in the sample, the fewer the amplification cycles required to detect it. This is why numerous studies on SARS-CoV-2 rely on *C*_*t*_ values to assess transmissibility (*2*) or study infection kinetics (*3*). Here, we present a cross-sectional analysis of SARS-CoV-2 RT-PCR tests performed on 2,220,212 individuals in France between January 21, 2020, and November 30, 2020 (Figure 1).

**Figure 1:**
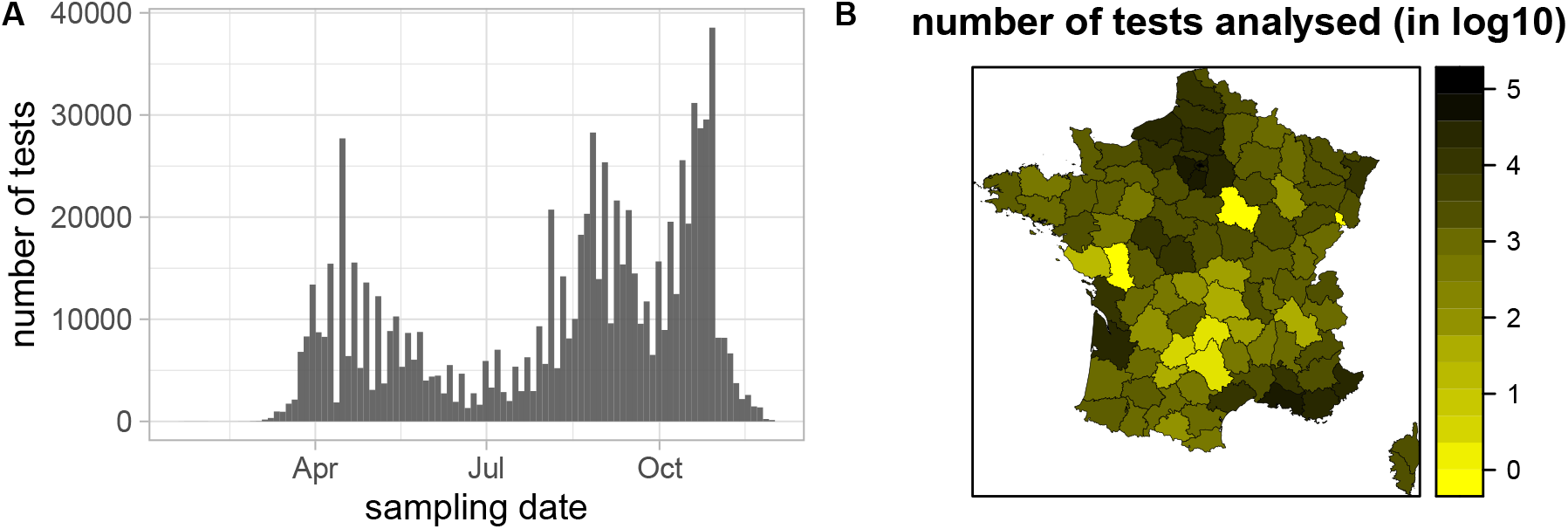
Number of tests performed per day (A) and per French department (B).

Few studies analyze *C*_*t*_ values at a population level. One explanation for this matter of affairs is that these are known to suffer from several, potentially strong, biases. First, sample type and sampling quality directly affect the amount of genetic material available. Second, the RT-PCR assay matters. Even the quality of the reagents used may have a significant effect on the number of amplification cycles required to achieve the same level of fluorescence for the same amount of target genetic material. Combining data from different laboratories helps to control for these sources of variation in statistical analyses. Furthermore, the larger the dataset, the more we can detect small statistical trends even after having controlled for non-informative variables.

In our analysis, we studied tests from individuals aged between 1 to 89 years old. We did not take into account tests for which key variables such as patient age, patient sex, laboratory geographical department, qualitative result, or RT-PCR assay used were unknown. Note that one test could provide more than one *C*_*t*_ value if containing probes targeting multiple viral genes. According to the national guidelines (*4*), it is recommended to focus on the most sensitive target to categorize levels of viral excretion. After removing the biologically unrealistic *C*_*t*_ values that were lower than 10 or larger than 45, the 95% confidence interval (95CI) of the remaining values was [16.89; 35.56] (Table S1). These correspond to 793, 479 tests from the same number of individuals.

We used a linear regression model to explore how *C*_*t*_ values can be explained by the following variables: patient age and sex, the number of days since the onset of the symptoms (if known), the clinical sampling site (if known), the sampling facility (if known), the RT-PCR assay used, the target gene, the test’s qualitative result, the sampling date, the temporal reproduction number of the epidemic at the sampling date (denoted *R*_*t*_ and estimated using the EpiEstim method (*5*)), and a control variable. The latter corresponds to the last digit of the patient anonymity number and is expected to be independent of the *C*_*t*_ value. We also included in the model an interaction term between sampling date and *R*_*t*_. For this analysis, we excluded *C*_*t*_ values from internal controls. Univariate analyses are extremely sensitive to heterogeneity in the dataset. For instance, the age distribution from patients sampled in aged care homes is different than that from city screening facilities, and analyzing the ‘sampling facility’ factor alone could yield misleading result. This is why the analysis used here is multivariate and considers all the factors listed above simultaneously.

Overall, the linear model explained 38.8% of the variance in *C*_*t*_ values, and the residuals were normally distributed (Figure S2A). All the factors except the temporal reproduction number were significantly associated with *C*_*t*_ values using a classical 5% p-value criterion in an analysis of variance (ANOVA) with type II sums of squares. Even for the control variable, the p-value was 0.013 and patients with final digits 1 and 3 had *C*_*t*_ values slightly lower (−0.19 and −0.17 cycles) than patients with a 0 final digit. We therefore set our significance thresholds to 5% of that of the control variables, *i*.*e*. 6.5 *×* 10^−4^ to analyze main effects (Table 1).

**Table 1:**
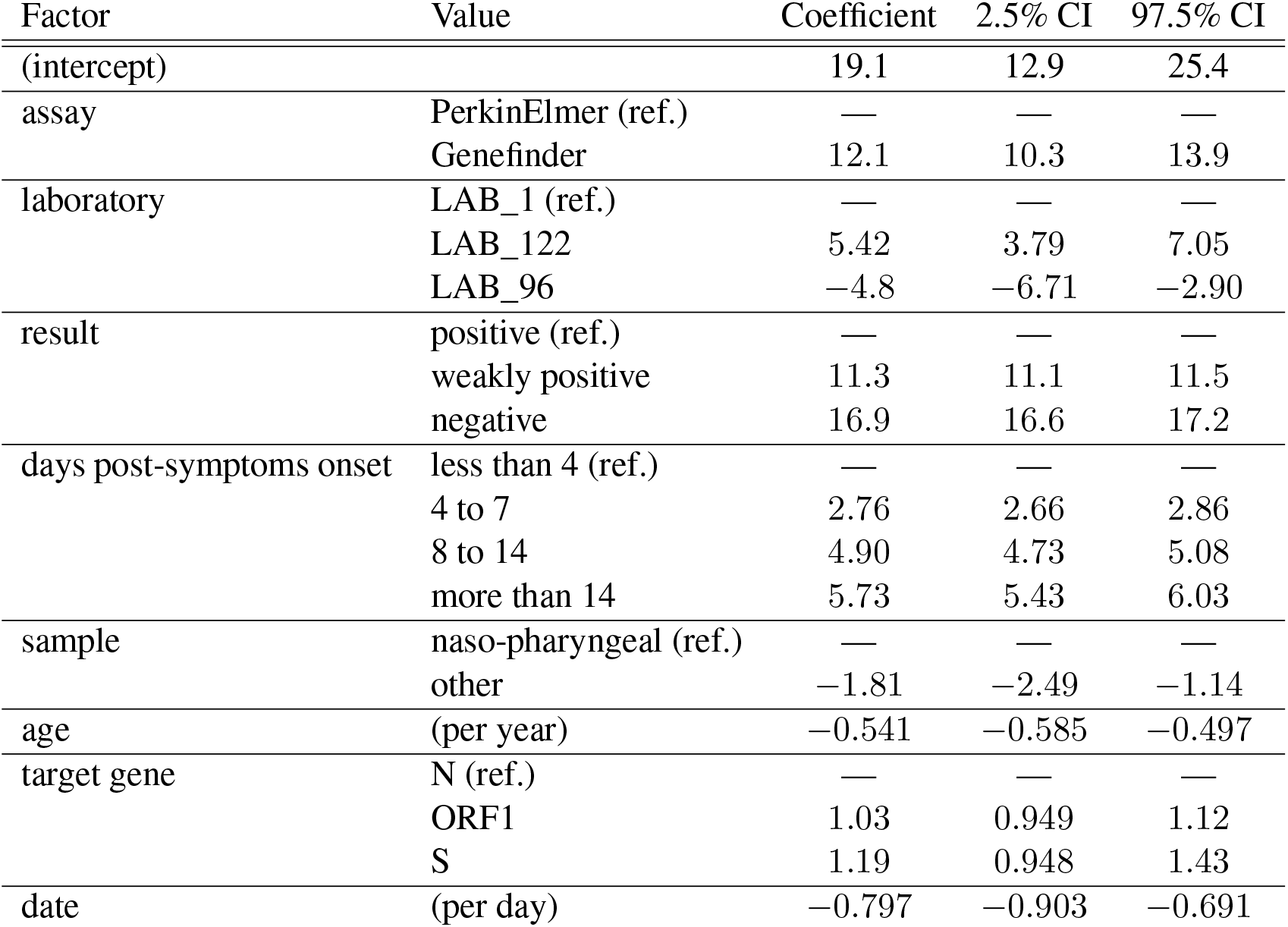
Main factors affecting Ct values. We only list factors with significant effects with a 10^−3^p-value criterion. Coefficients reflect differences in *C*_*t*_. For qualitative factors, the reference value is shown.

| Factor | Value | Coefficient | 2.5% CI | 97.5% CI |
| --- | --- | --- | --- | --- |
| (intercept) |  | 19.1 | 12.9 | 25.4 |
| assay | PerkinElmer (ref.) | — | — | — |
|  | Genefinder | 12.1 | 10.3 | 13.9 |
| laboratory | LAB_1 (ref.) | — | — | — |
|  | LAB_122 | 5.42 | 3.79 | 7.05 |
|  | LAB_96 | −4.8 | −6.71 | −2.90 |
| result | positive (ref.) | — | — | — |
|  | weakly positive | 11.3 | 11.1 | 11.5 |
|  | negative | 16.9 | 16.6 | 17.2 |
| days post-symptoms onset | less than 4 (ref.) | — | — | — |
|  | 4 to 7 | 2.76 | 2.66 | 2.86 |
|  | 8 to 14 | 4.90 | 4.73 | 5.08 |
|  | more than 14 | 5.73 | 5.43 | 6.03 |
| sample | naso-pharyngeal (ref.) | — | — | — |
|  | other | −1.81 | −2.49 | −1.14 |
| age | (per year) | −0.541 | −0.585 | −0.497 |
| target gene | N (ref.) | — | — | — |
|  | ORF1 | 1.03 | 0.949 | 1.12 |
|  | S | 1.19 | 0.948 | 1.43 |
| date | (per day) | −0.797 | −0.903 | −0.691 |

The intercept of the linear model indicates the average *C*_*t*_ value for a positive test performed with the reference assay, and all the other factors being set to their reference value. Its magnitude (19.1 cycles) is in line with clinical practice. The importance of the noise in the dataset is illustrated by the strong effect of the testing laboratory, as well as the RT-PCR assay used (Supplementary Figure S1).

Despite this high level of noise, we detected a strong effect of the qualitative result (Figure 2A), with *C*_*t*_ differences that were even larger than that from the laboratory effect. We also found a slightly significant difference of −1.81 cycles between the most common type of samples (nasopharyngeal) and that performed on other clinical sampling sites (mostly lower respiratory tracts, but also feces or saliva). This is likely because the latter tests were performed in patients with more severe symptoms.

**Figure 2:**
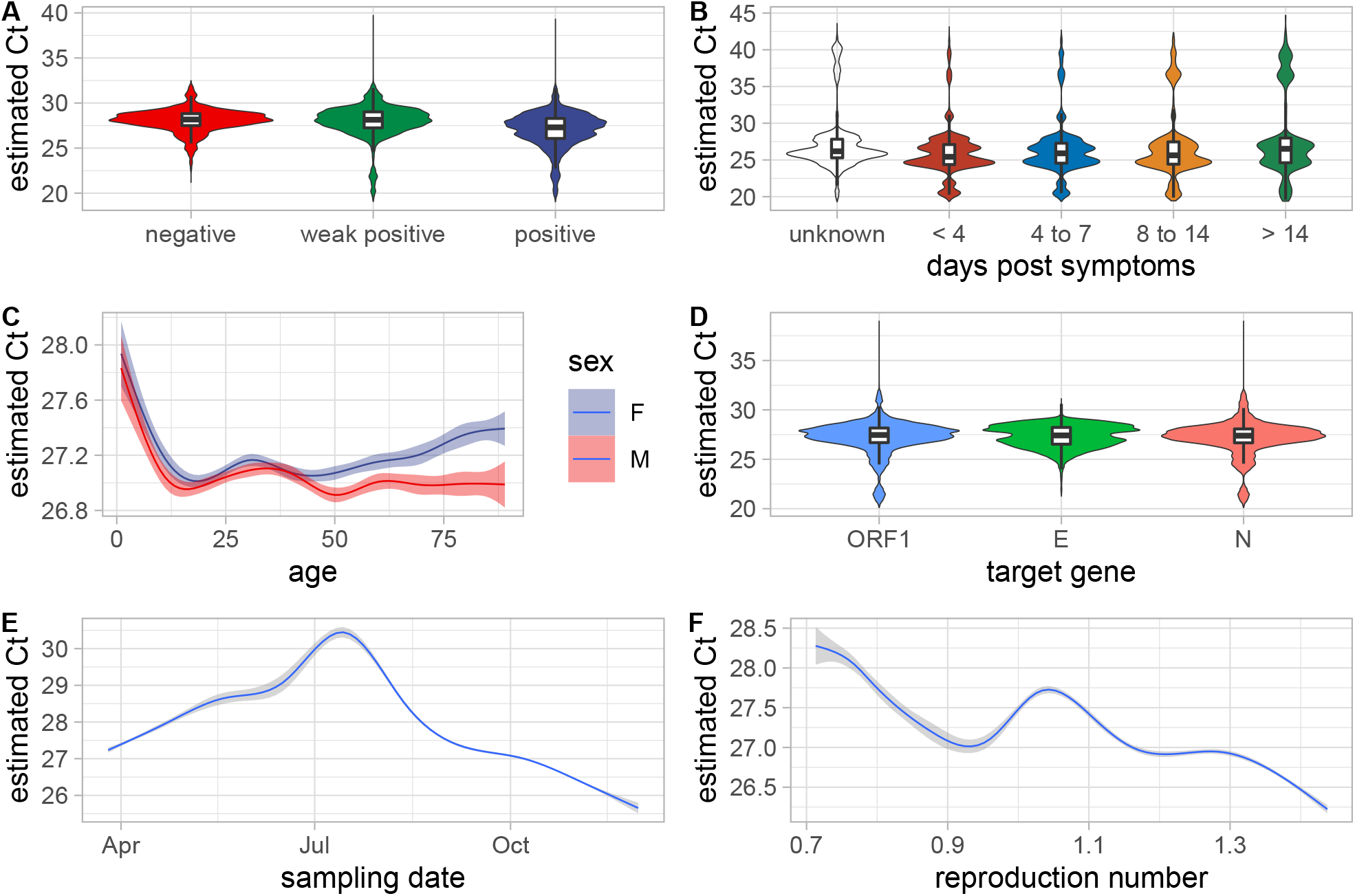
Correlations between key factors and observed *C*_*t*_ variations. A) Qualitative result of the test, B) number of days between symptoms onset and testing, C) participant age and sex, D) genomic area targeted by the test, E) sampling date, and F) temporal reproduction number (*R*_*t*_) at the time of the test. For panels A, B, and D, the violin plots indicate the distributions and the box plots show the 0.025, 0.25, 0.5, 0.75, and 0.975 quantiles. Panels C, E, and F are obtained with a ‘loess’ smoothing model. The *C*_*t*_ values shown are not the raw values but that estimated using a linear model to correct for biases (see Supplementary Methods for details).

The effects associated with the number of days since symptoms onset was particularly strong. For the 8.5% of the participants for whom the number of days between symptoms onset and testing dates was known, we found that the *C*_*t*_ gradually increases over the reported range with a maximum difference of 5.73 cycles (Figure 2B).

The effect of sex had the same order of magnitude as that of the control variable and could, therefore, be treated as non-significant. Conversely, the age factor had a strong effect with a decrease of 0.541 cycles per year (Figure 2C).

The target gene of the RT-PCR assay used also yielded a slightly significant effect. The *C*_*t*_ values obtained when using a probe targeting the ORF1 and S regions of the virus genome were significantly higher than when using the N gene, which was the genomic region of reference in the model (Figure 2D). This effect is consistent with the life-cycle of the virus. As stressed by (*6*), since coronaviruses are (+)ssRNA viruses, they use the same RNA matrix for replication and transcription, both being amplified by diagnostic assays. Furthermore, *Coronaviridae* transcripts can produce subgenomic mRNAs that lack part of the genome (*7*). As a consequence, and as shown in cell cultures (*8*), genes on the 5’ end of the genome are under-represented. This is consistent with our result where assays targeting the gene on the 3’ end (the N gene) tend to have lower *C*_*t*_ than assays targeting genes on the 5’ end (the ORF1 and S genes). Note that an alternative explanation could be that some probes target more conserved areas of the SARS-CoV-2 than others (*9*).

Finally, we found that *C*_*t*_ values decreased with time (−0.797 cycle per day), but this effect was non-linear (Figure 2E). This could be due to the strong variation in testing efforts in France (Figure 1A), but also to variations in the epidemic trend. Indeed, although the *R*_*t*_ (inferred from hospitalization data using the EpiEstim method (*5*)) was not found to be significant, the interaction between the sampling date and *R*_*t*_ was nearly significant (Figure 2F), suggesting that a temporal analysis could yield additional insights. The existence of a correlation between the *C*_*t*_ values of the tests performed in a population and *R*_*t*_ is consistent with population dynamics theory, which predicts that in an expanding population of infected individuals, the ‘age’ of the infections, *i*.*e*. the number of days post-infection, is skewed towards lower values (*10*). Since *C*_*t*_ values have been reported to increase over the course of an infection (*3*), which we confirm with this analysis (Figure 2D), it has suggested that these values could be used as an early signal to predict *R*_*t*_ (*11*).

To investigate this question, we focused on screening data collected in the general population from individuals aged from 5 to 79. We estimated the median and skewness values of the daily distribution of the *C*_*t*_ values. To correct for potential confounding factors, these were adjusted using a linear model (see the Supplementary Methods). We analyzed the temporal correlation between the time series with a 7-day rolling average of this median, skew, and *R*_*t*_ (Figure 3). For the median *C*_*t*_ value, we found a significant correlation with *R*_*t*_ that was maximized for a 6 to 7 days delay (Figure S2). This is consistent with *R*_*t*_ being calculated using data from ICU-admissions, which occur with a median of 14 days after infection (*12, 13*), and RT-PCR screening data being obtained earlier in the infection.

**Figure 3:**
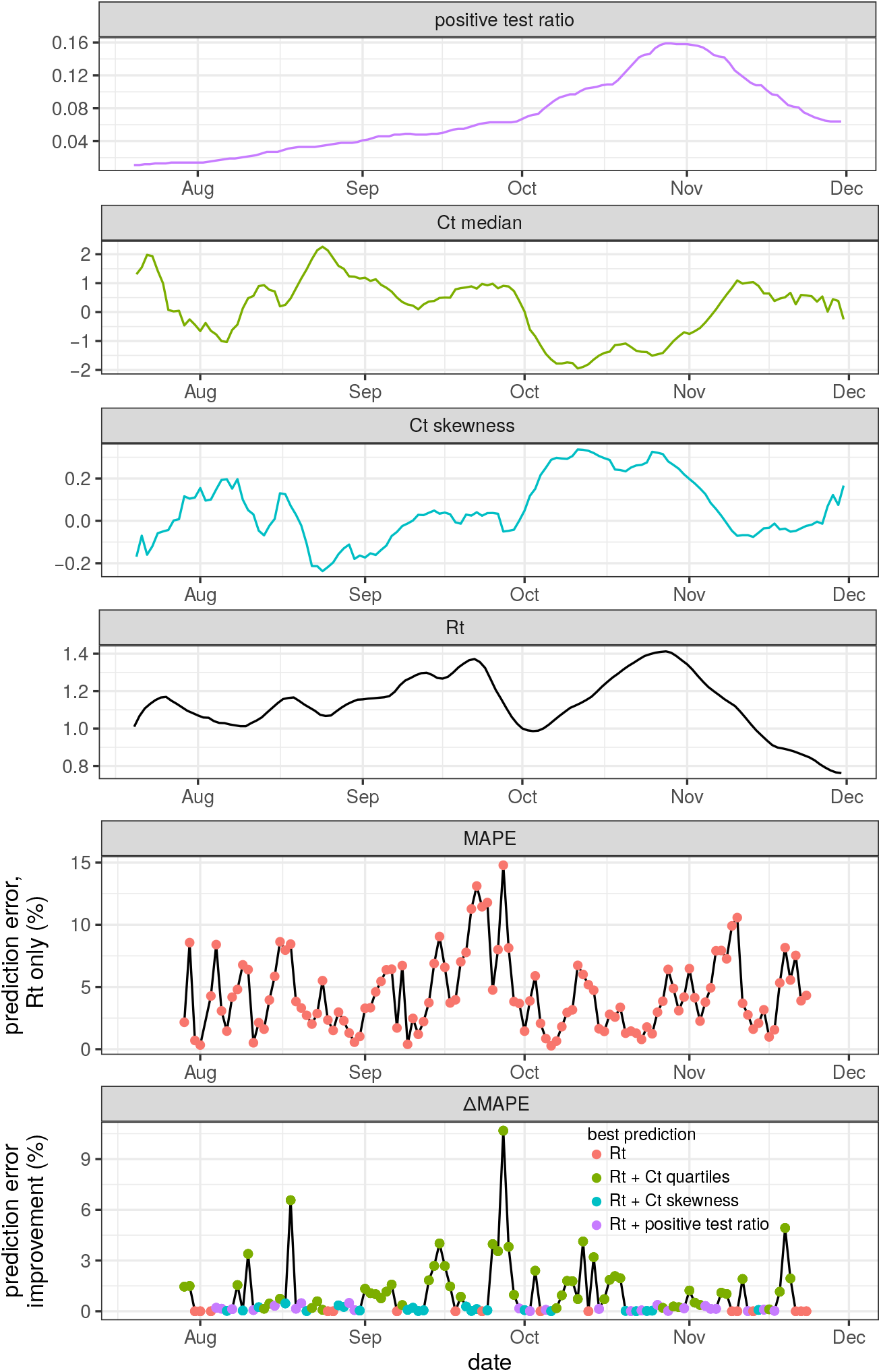
Predicting temporal reproduction number (*R*_*t*_) from time series. The top four panels show the the 7-days rolling averages of the time series of the ratio of positive tests (in purple), the median (in green) and skewness (in cyan) of the daily *C*_*t*_ residual distribution, and *R*_*t*_ (in black). The bottom panels show the error made by a prediction using only *R*_*t*_ data (red dots) and the potential improvement made by including exogeneous data.

To further assess the usefulness of *C*_*t*_ data, we used ARIMA models to predict *R*_*t*_ dynamics over 7 days. We compared models without any exogenous data, to models that also included exogenous time series (either median or skewness of estimated *C*_*t*_ values distribution, or the fraction of positive tests (*1*)). As expected, the prediction error made using only endogenous data (*R*_*t*_) was low in periods where *R*_*t*_ variations were limited. Furthermore, we found that adding exogenous data improved the prediction, especially when strong shifts in *R*_*t*_ were occurring (Figure 3). *C*_*t*_ values (green and cyan dots) tended to provide a better reduction in the error of the prediction than the ratio of positive tests (purple).

This analysis of a large national database of RT-PCR tests performed in the context of a major epidemic confirms that population-level *C*_*t*_ values are noisy since even a linear model that features 91 degrees of freedom does not explain the majority of the variance. However, owing to the law of large numbers, we detect several effects that are in line with biological observations and with virological properties. For instance, our finding that *C*_*t*_ values decrease as a function of the number of days after symptoms onset is consistent with longitudinal follow-ups (*3*). Similarly, the difference we detect between the virus gene targeted by the RT-PCR assay used can be interpreted in the light of known differences in mRNA copy numbers between genes depending on their distance from the 3’ end (*8*). Concerning the link between age and *C*_*t*_ values, although there are some mechanistic interpretations as to why virus load would increase with age (*14*), the evidence was mixed, with some studies reporting a decreasing trend (*15*) and others not (*16, 17*). Here, using a multivariate approach on a large dataset allows us to unravel a strong and significant decrease of *C*_*t*_ values with age.

A limitation our study is that although our dataset stands out by its size and its level of details, it is restricted to a single country where testing effort varies, both on a temporal and on a spatial scale (Figure 1). Performing similar analyses for other populations can therefore be particularly informative. A promising output of this analysis is the possibility to use *C*_*t*_ values as an early signal to detect changes in epidemic behavior, *e*.*g. R*_*t*_ values. Indeed, our most robust descriptors of the epidemics originate from hospital-admission data, but these have a significant delay with the status of the epidemic since patients are hospitalized 2 weeks after infection (*12,13*). The ratio of positive tests performed in the population of interest can, in theory, provide earlier insights but it suffers from strong sampling biases. We show that accounting for population-based *C*_*t*_ values variations can improve *R*_*t*_ predictions on a 7-days period. This corroborates earlier hypotheses (*11*) but also reveals the importance of analyzing a large-enough dataset to filter out the important amount of noise in these values. Overall, these results call for better integration of *C*_*t*_ values in national public health policies to monitor epidemics caused by SARS-CoV-2 but also other human viruses, especially since these data raise less ethical concerns than other sources of data such as mobility data.

## Data Availability

Data will be made available upon publication of the manuscript.

## Supplementary materials

### S1 Authors contributions

SA, VF, and SB conceived the study, SH-B, J-MG, LN, SP, TV, AdR, CT, MS, RS, CB-B, MS, A-SLH, SB, CL, JD, SH, CH, DV, HMD, SF, BV,CH, VF, and SB contributed anonymous data, SA compiled the anonymous data sets from the collaborating partners, SA, MTS, and CS analysed the data, SA wrote the first draft of the manuscript, all authors commented and approved the manuscript.

This study was supported by the COVID-19 study group from the Société Française de Microbiologie (SFM).

### S2 Ethics

This study was approved by the Internal Review Board of the Centre Hospitalier Universitaire de Montpellier. It is registered at ClinicalTrials.gov under the identifier NCT04738331.

### S3 Data and scripts

The final data set analyzed along with the R scripts will be made available upon publication.

### S4 SFM COVID-19 study group

The participants of the SFM COVID-19 study group are LINA Gérard (president of the SFM, CHU Lyon), VABRET Astrid (CHU de Caen), ADNET Justine (CHU de Caen), ROQUEBERT Benedicte (Cerba), DUCANCELLE Alexandra (CHU d’Angers), LE GUILLOU-GUILLEMETTE Hélène (CHU d’Angers), BOUTHRY Elise (CHU d’Angers), LUNEL-FABIANI Françoise (CHU d’Angers), PIVERT Adeline (CHU d’Angers), APAIRE-MARCHAIS Véronique (CHU d’Angers), ROGER Steven (CHU d’Angers), Chakib Alloui (Avicenne), Ségolène Brichler (Avicenne), Emmanuel Gordien (Avicenne), MIRAND Audrey (CHU de Clermont-Ferrand), ARCHIMBAUD Christine (CHU de Clermont-Ferrand), BREBION Amélie (CHU de Clermont-Ferrand), REGAGNON Christel (CHU de Clermont-Ferrand), CHABROLLES Hélène (CHU de Clermont-Ferrand), BISSEUX Maxime (CHU de Clermont-Ferrand), COMBES Patricia (CHU de Clermont-Ferrand), Hélène Jeulin (CHU de Nancy), Véronique Venard (CHU de Nancy), Evelyne Schvoerer (CHU de Nancy), Anne Lebouter (Henri Mondor APHP), Souraya Khouider (Henri Mondor APHP), Alexandre Soulier (Henri Mondor APHP), Aurelie Gourgeon (Henri Mondor APHP), Bellecave Pantxika (CHU de Bordeaux), Busson Laurent (CHU de Bordeaux), Garrigue Isabelle (CHU de Bordeaux), Lafon Marie-Edith (CHU de Bordeaux), Trimoulet Pascale (CHU de Bordeaux), Bruno Pozzetto (CHU de Saint-Etienne), Thomas Bourlet (CHU de Saint-Etienne), Sylvie Gonzalo (CHU de Saint-Etienne), Rémi Labetoulle (CHU de Saint-Etienne), Rogez Sylvie (CHU de Limoges), Alain Sophie (CHU de Limoges), Marianne Coste-Burel (CHU de Nantes), Virginie Ferré (CHU de Nantes), Berthe-Marie Imbert-Marcille (CHU de Nantes), Pierre Edouard Fournier (IHU Méditerranée infection), Petit Paul Rémi (IHU Méditerranée infection), Luciani Léa (IHU Méditerranée infection), Zandotti Christine (IHU Méditerranée infection), Charre Caroline (Cochin APHP), Mariaggi Alice-Andrée (Cochin APHP), Méritet Jean-François (Cochin APHP), Rozenberg Flore (Cochin APHP), Febreau Christine (CHU de Rennes), Comacle Pauline (CHU de Rennes), Lagathu Gisèle (CHU de Rennes), Maillard Anne (CHU de Rennes), Grolhier Claire (CHU de Rennes), Pronier Charlotte (CHU de Rennes), David Boutolleau (La Pitié Salpêtrière APHP), Anne-Geneviève Marcelin (La Pitié Salpêtrière APHP), Vincent Calvez (La Pitié Salpêtrière APHP), Stéphane Marot (La Pitié Salpêtrière APHP), Sepideh Akhavan (La Pitié Salpêtrière APHP), Basma Abdi (La Pitié Salpêtrière APHP), Marc Wirden (La Pitié Salpêtrière APHP), Cathia Soulié (La Pitié Salpêtrière APHP), Aude Jary (La Pitié Salpêtrière APHP), Elisa Teyssou (La Pitié Salpêtrière APHP), Sylvie van der Werf (CNR IPP), Vincent Enouf (CNR IPP), BOUDET Agathe (CHU de Nîmes), CARLES Marie-Josee (CHU de Nîmes), PERE Hélène (HEGP APHP), BELEC Laurent (HEGP APHP), IZQUIERDO Laure (HEGP APHP), RODARY Julien (HEGP APHP), BAILLARD Jean-Louis (HEGP APHP), RIBEYRE Tatiana (HEGP APHP), SALIBA Madelina (HEGP APHP), ROGER Alicia (HEGP APHP), GARNIER Nathalie (HEGP APHP), ROBILLARD Nicolas (HEGP APHP), Le Goff Jérôme (Saint Louis APHP), Delaugerre Constance (Saint Louis APHP), Chaix Marie-Laure (Saint Louis APHP), Feghoul Linda (Saint Louis APHP), Mahjoub Nadia (Saint Louis APHP), Maylin Sarah (Saint Louis APHP), Schnepf Nathalie (Saint Louis APHP), Alfaisal Jamal (Saint Louis APHP), AGNELLO Davide (CHU de Dijon), AUVRAY Christelle (CHU de Dijon), BELLIOT Gaël (CHU de Dijon), BOUR Jean-Baptiste (CHU de Dijon), CASENAZ Alice (CHU de Dijon), GUILLOTIN Florence (CHU de Dijon), MANOHA Catherine (CHU de Dijon), SI-MOHAMMED Ali (CHU de Dijon), TAN Rithy-Nicolas (CHU de Dijon), Diane Descamps (Bichat APHP), Nadhira Houhou-Fidouh (Bichat APHP), Charlotte Charpentier (Bichat APHP), Houria Ichou (Bichat APHP), Florence Damond (Bichat APHP), Quentin Le Hingrat (Bichat APHP), Valentine Ferré (Bichat APHP), Lucile Larrouy (Bichat APHP), Vincent Mackiewicz (Bichat APHP), Gilles Collin (Bichat APHP), FAFI-KREMER Samira (CHU de Strasbourg), GALLAIS Floriane (CHU de Strasbourg), LAUGEL Elodie (CHU de Strasbourg), BENOTMANE Ilies (CHU de Strasbourg), VELAY Aurélie (CHU de Strasbourg), and WENDLING Marie-Josée (CHU de Strasbourg)

## S5 Supplementary Methods

### S5.1 Initial data filtering

The French Society for Microbiology (SFM) send a query to collect anonymous RT-PCR test results data from 19 public and 2 private laboratories. The response files were curated manually and merged using R. Test values without any *C*_*t*_ values (negative tests) were removed.

This led to an initial global data set from 2, 220, 212 individuals. Removing non-numerical *C*_*t*_ values (usually a qualitative description of a negative result) decreased this number by 30% from 10, 668, 371 to 7, 516, 936 *C*_*t*_ values (note that most tests are usually associated with more than a single value since it can have multiple targets). We then removed all the *C*_*t*_ values equal to zero, which left us with 1, 969, 043 values.

Finally, we performed some extra filtering to remove aberrant *C*_*t*_ values (greater than 100), test with missing values for sampling French department, qualitative result, or RT-PCR test used. This left us with 1, 299, 447 values originating from 824, 446 individuals. Further details are available in Table S1.

For each individual, we retained only the earliest sample therefore analyzing 1, 129, 437 *C*_*t*_ values.

### S5.2 Database properties

**Table S1:** Description of the dataset variables. For real numbers, we show the median and the 95% confidence interval. For categorical variables, we either indicate the number of factors or the number of occurrences *n* for each factor.

| Variable | Description | Details | Values |
| --- | --- | --- | --- |
| department | French administrative department where the sampling was performed | categorical | 97 departments |
| id_lab | laboratory associated with the sampling | categorical | 128 labs |
| control_variable | a control variable created using the last digit of the patient anonymity number | categorical | 10 values |
| id_patient | participant anonymity number | categorical | 825, 446 ids |
| date | sampling date | date | min = 01/21/2020<br>max = 30/11/2020 |
| sampling_facility | type of facility where the sampling was performed | *city screening*<br>*aged care home*<br>*hospital*<br>*prison*<br>*missing* | $n = 1,008,307$<br>$n = 3,822$<br>$n = 72,682$<br>$n = 45$<br>$n = 44,570$ |
| sample_type | clinical localization sampled | *nasopharyngeal*<br>*other* | $n = 1,086,004$<br>$n = 35,660$ |
| assay_PCR | the description of the RT-PCR assay used | categorical | 13 assays |
| target_gene | SARS-CoV-2 genomic area corresponding to the $C_t$ value | $N$<br>$E$<br>$S$<br>$ORF1$<br>$N \& ORF1$<br>*internal control* | $n = 199,256$<br>$n = 55,717$<br>$n = 9,953$<br>$n = 227,192$<br>$n = 1,196$<br>$n = 636,123$ |
| $C_t$ | amplification cycles required to reach the signal (0 values are removed) | real | 28.10 [18.30; 37.37] |
| $R_t$ | temporal reproduction number (see Supplementary methods for details) | real | 1.15 [0.76; 1.41] |
| result | qualitative result of the test | *positive*<br>*negative*<br>*weak positive* | $n = 524,065$<br>$n = 585,293$<br>$n = 20,079$ |
| age | participant age (in years) | integer | 48 [25; 83] |
| sex | patient sex | *female*<br>*male* | $n = 637,212$<br>$n = 492,225$ |
| symptoms | number of days between symptoms onset and testing | *less than 4*<br>*4 to 7*<br>*8 to 12*<br>*more than 12*<br>*missing* | $n = 107,417$<br>$n = 22,308$<br>$n = 6,070$<br>$n = 1,945$<br>$n = 991,697$ |

### S5.3 Temporal reproduction number (*R*_*t*_)

The temporal reproduction number (denoted *R*_*t*_) was computed on the COVID-19 hospital admission incidence time series established by the national public health agency (Santé Publique France) and accessible at this website. Because of strong daily variations (especially on week-ends), we first transformed the time series using a 7-days rolling average. We then used the EpiEstim method (*5*) and the eponym package in R (*18*).

Earlier studies have reported that, for patients who develop severe symptoms, the median time between infection and hospital admission is in the order of 14 days (*12, 13*).

### S5.4 Statistical analyses

All the analyses were performed in R version 3.6.3.

#### S5.4.1 Linear model

The linear model was performed in individuals from 1 to 90 years old. We also removed *C*_*t*_ values associated to internal controls because they caused the distribution of the residuals to be non-Gaussian. Quantitative factors, namely *R*(*t*), date, and age were scaled and centered.

The model was formulated as follows in R:

~~~
modele_general = lm(Ct ∼ Rt*date + age + sexe + target_gene +
assay_PCR + id_lab + symptoms + result +
sample_type + sampling_facility + control_variable, data)
~~~

The adjusted *R*^2^ of the model was 38.8% and the distribution of the residuals Gaussian (Figure S2A). The model was analyzed using an ANOVA assuming type-II errors because of the unbalanced nature of the data set. All the variable were found to be extremely significant (p-value *<* 10^−6^), except for *R*_*t*_ (p-value of 0.68) the control variable (p-value of 0.0131), sampling facility (p-value of 0.0135), sex (p-value of 0.0021), interaction between *R*_*t*_ and date (p-value of 0.000842).

#### S5.4.2 Time series analyses

To analyze the time series of reproduction number and *C*_*t*_ values we restricted the data to tests performed after July 1, 2021, in a screening context, using nasopharyngeal swabs, in individuals aged from 6 to 80. These assumptions were made such that *C*_*t*_ values would reflect the state of the ongoing epidemic. Finally, we excluded values from internal control genes. Overall, we analysed 234,782 *C*_*t*_ values from 110,227 individuals.

To correct for other potential confounding factors, we first performed a linear model.

The analyses were performed on the residuals of the linear model. For each day, we computed the median and the skewness of the distribution. Skewness was computed using the formula 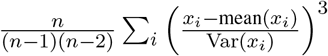 with *x*_*i*_ the individual values and *n* the total number of values. We then computed a 7-day rolling mean to buffer daily variations.

The temporal reproduction number was computed as indicated above. We also analyzed a 7-day rolling mean.

Cross-correlation function analyses were performed using the ccf function in R.

#### S5.4.3 Predictive analyses

We used ARIMA models (implemented in the R package forecast) to predict the hospital-admission temporal reproduction number (*R*_*t*_) from past observations.

For each date, predictions were evaluated in terms of the mean absolute percentage error (MAPE) for horizons of 7 days in the future based on coefficients learned from past data starting on July 29th, 2020. More precisely, for each date we compared the temporal reproduction number data {*D*_*k*_; *k* = 1, …, 7} with the seven-day model forecast {*F*_*k*_; *k* = 1, …, 7} by

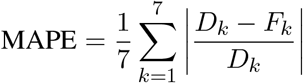

We considered 4 types of data:

1. past *R*_*t*_ data (i.e. endogenous data),
2. quartiles from *C*_*t*_ residuals (to remove biases),
3. skewness from Ct residuals,
4. national positive test ratio collected from https://covid.ourworldindata.org/data/.

The residuals of *C*_*t*_ were obtained from the following linear model:

~~~
lm(Ct ∼ age + target_gene*assay_PCR + id_lab, data)
~~~

Our goal was to see if adding exogenous data, i.e. *C*_*t*_ values and proportion of positive tests, increased prediction precision.

Models were tuned with the auto.arima function, and untuned models were run with *p* = 9, *d* = 2, and *q* = 0 as default parameters, based on the cross-correlation analysis between *R*_*t*_ and *C*_*t*_ time series (Figure S2).

Prediction improvement by models using in addition exogenous data relative to model using past values of *R*_*t*_ only was defined by

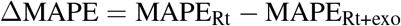

##### S5.4.4 Figure 2 *C*_*t*_ values

To visualize the effect of the different factors, we first perform a linear model to correct for the effect of confounding factors.

We used the general model described above but removed the effect ‘control_variable’, as well as the effects ‘target_gene’, ‘sample_type’, ‘symptoms’ and ‘sampling_facility’ which had little effect and were sometimes lacking for many participants. For each figure, we also removed the main factor of interest (depending on the panel).

The *C*_*t*_ value itself was generated using the predict.lm function in R.

### S6 Supplementary Figures

**Figure S1:**
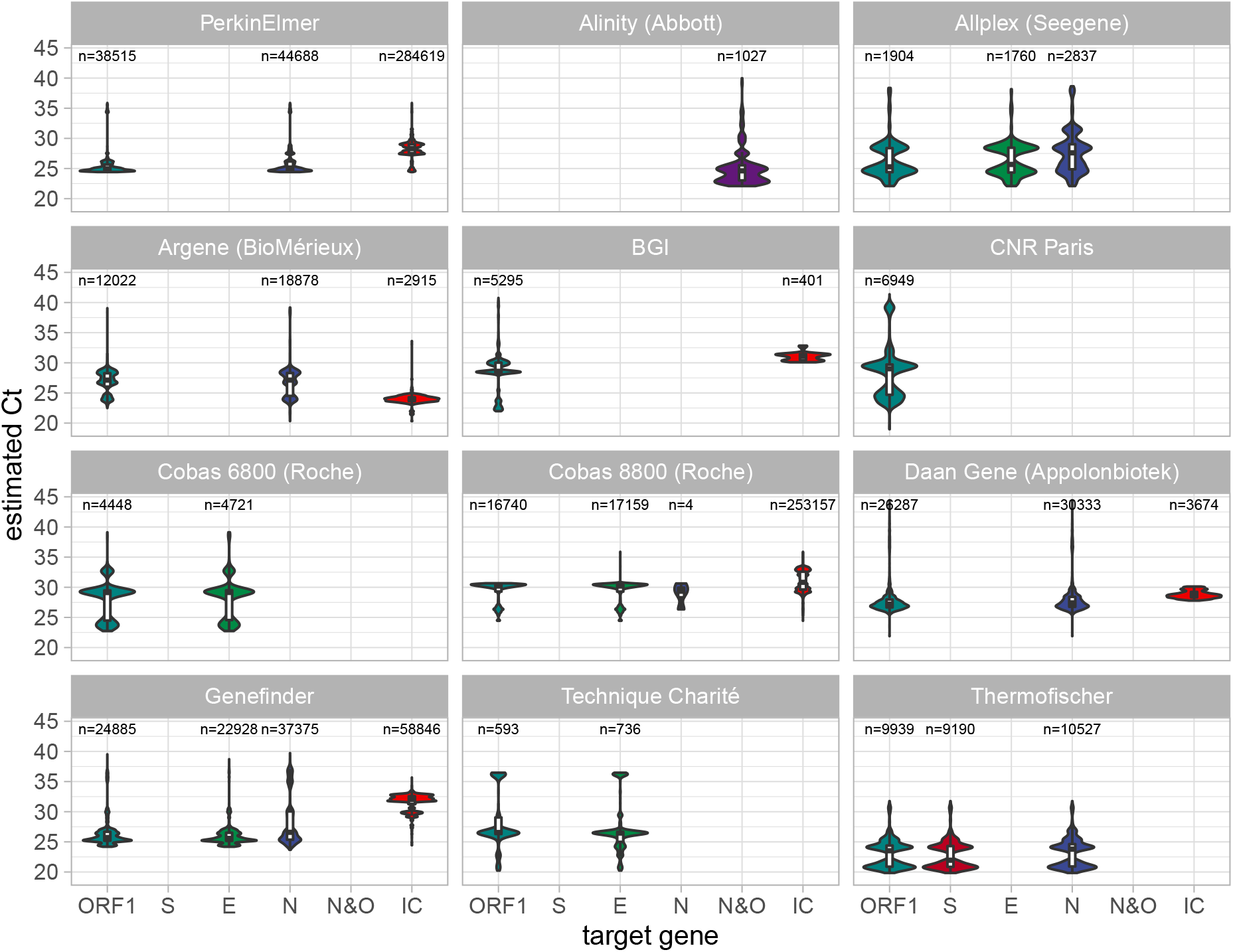
Effect of the RT-PCR assay used on the estimated *C*_*t*_ value as a function of the targeted genomic area. Only tests with at least 1,000 *C*_*t*_ values are shown.

**Figure S2:**
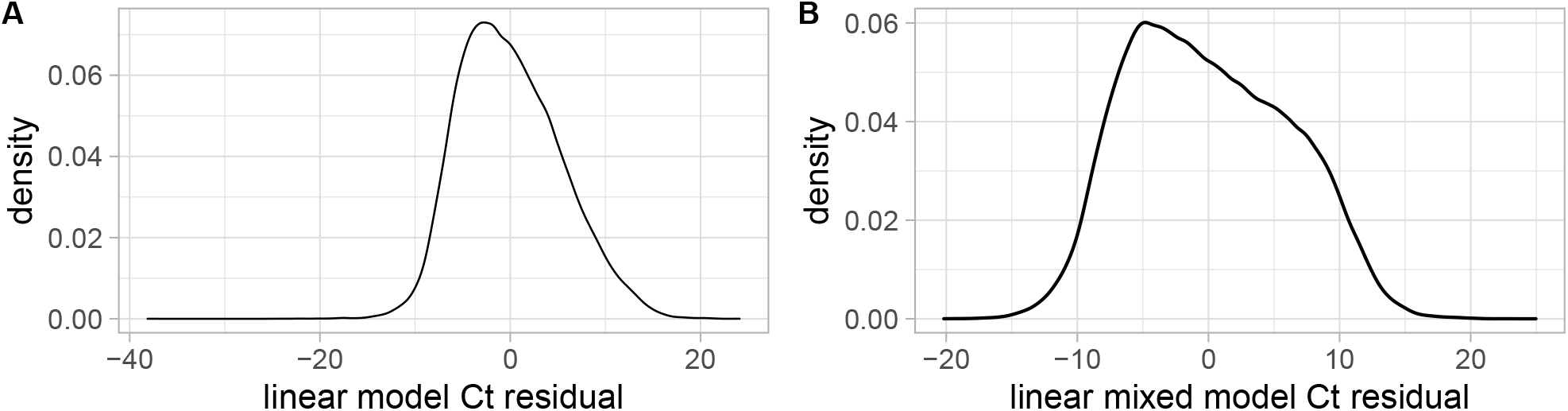
Distribution of the *C*_*t*_ residuals value. A) For the linear model used for the main analysis shown in Table 1, B) for the linear model used to generate residuals for the *R*_*t*_ time series analysis.

**Figure S3:**
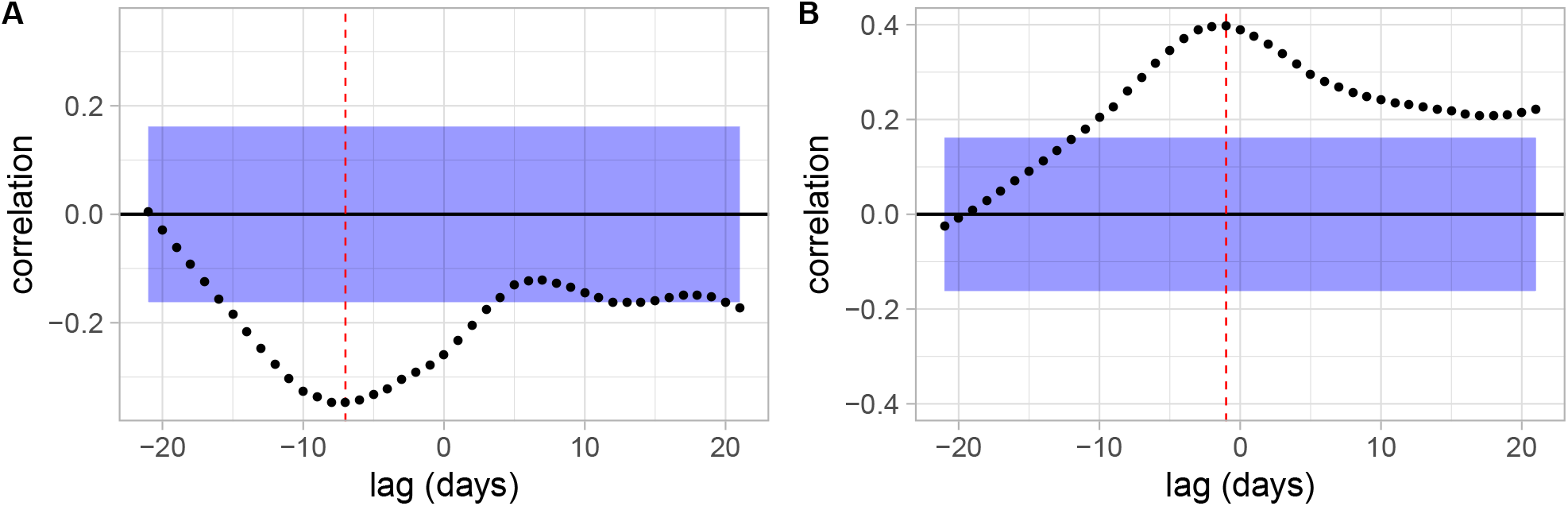
Cross correlation functions between *R*_*t*_ and A) the median or B) the skewness of the *C*_*t*_ residuals distribution. The blue shaded areas show the non-significant values (with a 95% threshold) and the red vertical dotted lines the lag with the highest significant correlation. Note that the lag is smaller for the skewness than for the median of the distribution.

**Figure S4:**
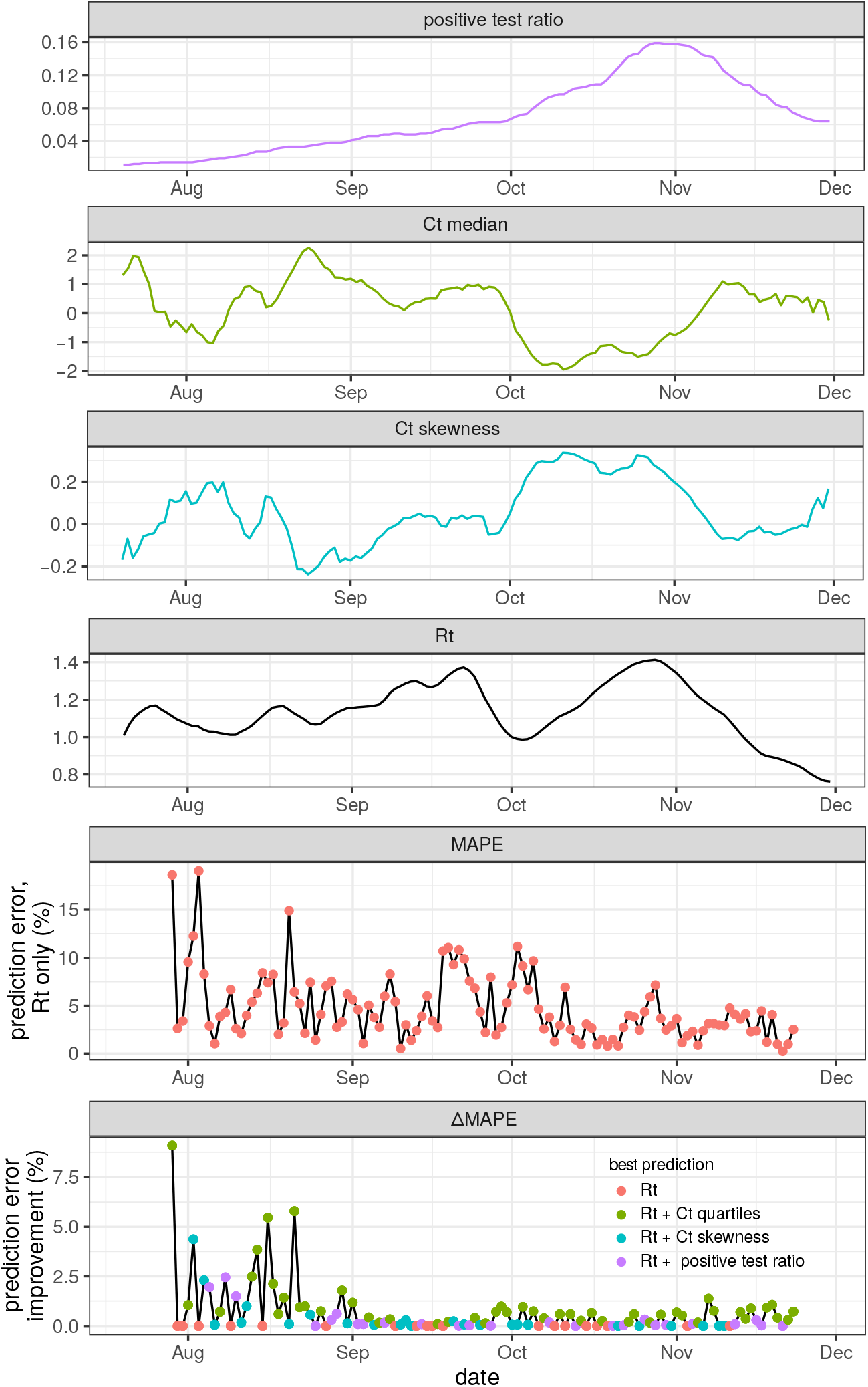
Predicting temporal reproduction number (*R*_*t*_) from time time series with untuned ARIMA parameters. *p* = 9, *d* = 2, *q* = 0 The top four panels show the the 7-days rolling averages of the time series of the ratio of positive tests (in purple), the median (in green) and skewness (in cyan) of the daily *C*_*t*_ residuals distribution, and *R*_*t*_ (in black). The bottom panels show the error made by a prediction using only *R*_*t*_ data (red dots) and the potential improvement made by including exogeneous data.

